# Cognitive function and vitamin B12 and D in elders from Ecuador

**DOI:** 10.1101/2021.01.17.21249997

**Authors:** Andrés Fernando Vinueza Veloz, Tannia Valeria Carpio Arias, Jenifer Sthefania Vargas Mejía, Estephany Carolina Tapia Veloz, Jefferson Santigo Piedra Andrade, Tomás Marcelo Nicolalde Cifuentes, Susana Isabel Heredia Aguirre, María Fernanda Vinueza Veloz

## Abstract

**Introduction:** Current evidence still does not support the role of vitamin B12 or vitamin D in age-associated cognitive impairment.

**Objective:** Evaluate the association between vitamin B12 and D and cognitive function in elders.

**Methodology:** Sample included 1733 individuals 60 years old and older, who participated in the SABE study that was carried out in Ecuador. Cognitive function was measured using abbreviated version of minimental state examination (MMSE). Vitamin B12 and D were measured in blood. Data were analyzed through linear regression models with restricted cubic splines (RCSs). Models were adjusted by sex, years of education, region (urban highland, urban coast, rural highlands, rural coast), socioeconomic status, and ethnicity.

**Results:** Independently from sex, age, years of education, ethnicity, socioeconomic status and geographical zone of residence, we found that vitamin B12 but not vitamin D levels were associated with cognitive function in a representative group of Ecuadorian elders. Elders with lower levels of vitamin B12 showed lower MMSE scores in comparison to elders with higher levels of vitamin B12. Moreover, a statically significant non linear interaction was found between vitamin B12 and age with respect to cognitive function. In this way, we observed that in elders 75 years old and older whose levels of vitamin B12 were 271 pg/ml or less, the drop of cognitive function was particularly steep in comparison to elders whose levels of vitamin B12 were 647 pg/ml or more.

**Conclusions:** Low levels of vitamin B12 but not of vitamin D are associated with low cognitive functioning.

## 1. Introduction

Cognitive abilities can be divided into domains, which include attention, memory, executive function, language and visuospatial abilities, among others. Each of these domains declines at different rates with age.(1) Specifically, studies in healthy older adults show declines in some cognitive domains including, attention, working memory and episodic memory in comparison to younger adults.(2) Cognitive impairment is a major public health problem, as it is one of the leading causes of disability and adverse health consequences in elder adults.(3) Given the current lack of therapies for cognitive impairment, actual efforts are focused on trying to identify factors that may influence its development and progression.(4)

Vitamin B12 also know as cobalamin, a water-soluble essential vitamin is one of the eight B vitamins. Vitamin B12 acts either as cofactor or coenzyme in a number of essential biochemical processes including DNA synthesis, cell metabolism and myelin maintenance.(5) In humans vitamin B12 is acquired from diet specially from meat, fish, shellfish and dairy products.(5) Vitamin D on the other hand is a fat-soluble secosteroid involved not only in calcium and phosphorus metabolism but also in genetic transcription via vitamin D receptors (VDR).(6,7) Since few foods contain vitamin D (e.g. oil fish, egg yolk, shiitake mushrooms, liver or organ meats), dermal synthesis after ultraviolet B ray exposition remains the major route to obtain vitamin D.(6)

During the last years, interest has risen around vitamins as key players in the prevention and treatment of cognitive decline.(8,9) In fact, associations between vitamin B12 or vitamin D and cognitive function in elders have been extensively investigated.(10,11) However, up to date evidence still does not support supplementation of vitamin B12 or vitamin D to prevent or treat cognitive decline.(12,13)In relation to normal aging, it is more likely that mild or sub-clinical vitamin deficiencies may play a role in cognitive decline rather than an overt vitamin deficiency.[14]Thus it is necessary to consider the status of vitamin B12 and vitamin D over a range of values and not just focus on deficiency when looking at cognitive outcomes.(5) The objective of the present work is to study the association of vitamin B12 and vitamin D values and cognitive function in a population-based context.

## 2. Methodology

### 2.1 Study design and setting

The present work is a cross-sectional study. Sample included elders who participated in the study ‥Salud, Bienestar y Envejecimiento‥ (SABE), which was carried out by the ‥ Ministerio de Inclusión Económica y Social‥ (MIES) and the ‥ Instituto Nacional de Estadísticas y Censos‥ (INEC) in Ecuador between 2009 and 2010. The objective of SABE was to evaluate health status, well-being, nutrition, family, work, and cognition of elders in Ecuador.(15) Data and operational manuals of SABE are publicly available and can be downloaded from INEC web page: https://anda.inec.gob.ec/anda/index.php/catalog/292

### 2.2 Participants of SABE

Participants of SABE came from urban and rural areas of the Littoral and Andes Mountains of Ecuador. Participants were selected by stratified, double stage probabilistic sampling performed by clusters so called domains (urban and rural areas from the Littoral and Andes Mountains) and proportional to the size of the population.(15) Strata refer to socioeconomic status (high, medium, low). During the first stage censal sectors were randomly selected from each domain. During the second stage dwellings were randomly selected form censal sectors and finally all elders were selected from the dwellings. SABE was carried out in two phases: during the first one, health and epidemiological data were collected (SABE I). During the second phase a sub-sample of the participants underwent biochemical evaluation (SABE II).(15)

### 2.3 Sample

Sample included all individuals from SABE II that is, all individuals who underwent biochemical evaluation (n = 2371). From them we randomly selected one individual from each dwelling (n = 1993). We excluded individuals who did not have data on their cognitive function (n = 160), whose levels of vitamin D or B12 were not registered (n = 20) or whose socio-demographic data (e.g age, ethnicity, years of education) were not available (n = 80). Thus, the final sample included 1733 male and female elders.

### 2.3 Variables

#### 2.3.1 Cognitive function

Cognitive function was evaluated using an abbreviated version of the Mini Mental State Examination (MMSE).(15,16) Score ranged from 0 to 19.

#### 2.3.2 Vitamin B12

Levels of vitamin B12 in pg/ml in blood were measured by electrochemiluminescence (Immulite 2000 Roche) following international recommendations.(15)

#### 2.3.3 Vitamin D

Levels of vitamin D as 25(OH)D in ng/ml in blood were measured by chemiluminescence (Hitachi, Roche Cobas e 411) following international recommendations. The lowest limit of detection for serum 25(OH)D assay was 4ng/ml.(15)

#### 2.3.4 Ethnicity

Ethnic group was determined by asking the participants: how do you identify yourself? Participants could identify themselves as ‘‘mestizo’’ (mixed ethnic ancestry), ‘‘indígena’’ (indigenous people), ‘‘blanco’’ (white) or another ethnic group (e.g black, mulato, other).(15)

#### 2.3.5 Socioeconomic status

Socioeconomic status was measured using an index that was developed applying principal component analysis. The index was constructed based on several variables including: health insurance and retirement, goods, housing quality, income and social subsidy.(15) The final variable includes 5 categories from 1 to 5. The lower the number the lower the socioeconomic status.

### 2.4 Statistical analysis

We considered MMSE as the main outcome variable. To model the outcome we considered two explanatory variables (vitamin B12 and vitamin D) and five covariates (sex [males vs. female], age in years, years of education, ethnicity, and region [urban highlands, urban coast, rural highlands, rural coast]). To investigate the association between MMSE and vitamin B12 and vitamin D we implemented linear regression models with restricted cubic splines (RCSs). Implemented RCS regression models determined the shape of the relationship between vitamin B12 and D and outcomes without any prior assumption. RCSs fitted a smooth continuous curve of adjusted means with 95% confidence intervals (95% CIs) across vitamin B12 and vitamin D levels. RCSs allowed for changes in the function at defined knot points and restricted the splines to linear relationships at the tail ends. Knot points were located at percentiles 5, 27.5, 50, 72.5, and 95, as previously recommended to avoid forcing curvature or inflections.(17)

All models were adjusted by the potential covariates listed above. Splines were applied to vitamin B12 and D, age, years of education; however, vitamin B12 and D did no show non-linear trends (both *p* > 0.05), thus splines were removed for these two variables. We also included interaction terms for vitamin B12 * age, vitamin B12 * ethnicity, vitamin B12 * sex, vitamin B12 * region, vitamin D * age, vitamin D * ethnicity, vitamin D * sex, and vitamin D * region. However, with the exception of vitamin B12 * age and vitamin D * age, there were no statistically significant interactions, and so non-statistically significant interaction terms were dropped from the model. Models were adjusted for sampling weights, which were extracted from SABE. Post-stratification weighting method was applied. Goodness-of-fit plots showed that chosen models fitted well with the data (Supplementary Figure S1). All statistical analyses were performed using R, RStudio and related packages available in R, including rms.(17–19)

### 3.5 Ethical considerations

The study was conducted following ethical standards established in Helsinki Declaration 1964.[15]Data of the study are publicly available and can be downloaded from the INEC web page.

## 3. Results

### 3.1 General characteristics of the sample

Final sample included 1733 elders, from whom 936 (54.01%) were female. Females were older than males and had less years of education. Distribution of ethnic groups was similar among females and males, with mestizo being the the most numerous group (Table 1). Distribution of SES among females and males shows that more than half of elders suffer from poverty. Distribution of region among females and males shows that more than half of elders lived in urban areas of the highlands and coastal region (Table 1).

**Table 1.** General characteristics of the sample. Sample included 1733 individuals. Abbreviations: IQR, interquartil range; n, number; %, percentage; SEE, socioeconomic status

|  |  | Female 936 (54.01%) |  | Male 797 (45.99%) |  |
| --- | --- | --- | --- | --- | --- |
|  |  | Median (IQR) | n (%) | Median (IQR) | n (%) |
| Age |  | 69 (64,76) |  | 68 (64,76) |  |
| Years of education |  | 3 (0,6) |  | 4 (2,6) |  |
| Ethnicity | Mestizo |  | 656 (70.09) |  | 568 (71.27) |
|  | Indigenous |  | 71 (7.59) |  | 62 (7.78) |
|  | White |  | 135 (14.42) |  | 86 (10.79) |
|  | Other |  | 74 (7.91) |  | 81 (10.16) |
| SES | 1 |  | 281 (30.02) |  | 235 (29.49) |
|  | 2 |  | 250 (26.71) |  | 179 (22.46) |
|  | 3 |  | 144 (15.38) |  | 169 (21.2) |
|  | 4 |  | 201 (21.47) |  | 126 (15.81) |
|  | 5 |  | 60 (6.41) |  | 88 (11.04) |
| Region | Urban highlands |  | 282 (30.13) |  | 209 (26.22) |
|  | Urban coast |  | 367 (39.21) |  | 285 (35.76) |
|  | Rural highlands |  | 188 (20.09) |  | 164 (20.58) |
|  | Rural coast |  | 99 (10.58) |  | 139 (17.44) |

Mean MMSE score for the sample was 14.50 (SD = 3.07), ranging from 4 to 19, with a median of 15. Mean vitamin B12 of the sample was 482.80 pg/ml, ranging from 149 to 1001, with a median of 414. Mean vitamin D of the sample was 26.36 ng/ml, ranging from 3 to 71.17, with a median of 25.28. According to guidelines, in general sample showed normal levels of vitamin B12, while insufficient levels of vitamin D.(20,21) Still, 38.49% (n = 667) of the sample showed deficiency of vitamin B12, being males more often affected (Table 2). It was also observed that females showed lower median MMSE score, as well as lower levels of vitamin D than males. However, insufficiency and deficiency of vitamin D occurred more often among males (Tabla 2).

**Table 2.** MMSE, vitamin B12 and vitamin D by sex. Abbreviations: IQR, interquartil range; MMSE, mini mental state examination

|  |  | Female |  | Male |  |
| --- | --- | --- | --- | --- | --- |
|  |  | Median (IQR) | n (%) | Median (IQR) | n (%) |
| MMSE |  | 14 (12, 16) |  | 15 (13, 17) |  |
|  |  | 480 (313.75, 718.25) |  | 360 (243, 539) |  |
| Vitamin B12 (pg/ml) | Normal (> 350) |  | 650 (69.44) |  | 416 (52.20) |
| | Deficiency (≤ 350) | | 286 (30.56) | | 381 (47.80) |
|  |  | 22.91 (18.29, 27.97) |  | 28 (23.02, 33.82) |  |
| Vitamin D (ng/ml) | Normal (> 30) |  | 322 (37.53) |  | 107 (14.40) |
|  | Insufficiency (21-29) |  | 362 (42.19) |  | 312 (42.00) |
|  | Deficiency (< 20) |  | 174 (20.28) |  | 324 (43.60) |

### 3.2 Cognitive function and vitamin B12 and vitamin D

Vitamin B12, but not vitamin D was statically significantly associated with cognitive function [vitamin B12: F(5) = 4.21, *p* = 0.001; vitamin D: F(5) = 0.76, *p* = 0.580]. In order to understand the effect of vitamin B12 on cognitive function we compared predicted MMSE scores among the first (271 pg/ml) and the third quartil (647 pg/ml) of vitamin D. We observed that MMSE scores were higher for elders who had highest vitamin B12 values (Figure 1, panel a). Moreover, a statically significant non linear interaction was found between vitamin B12 and age with respect to cognitive function [F(3) = 3.93, p = 0.008]. In this way, we observed that in elders 75 years old and older whose levels of vitamin B12 were 271 pg/ml or less, the drop of cognitive function was particularly steep in comparison to elders whose levels of vitamin B12 were 647 pg/ml or more. Such an effect was not observed in the case of vitamin D (Figure 1, panel a and b).

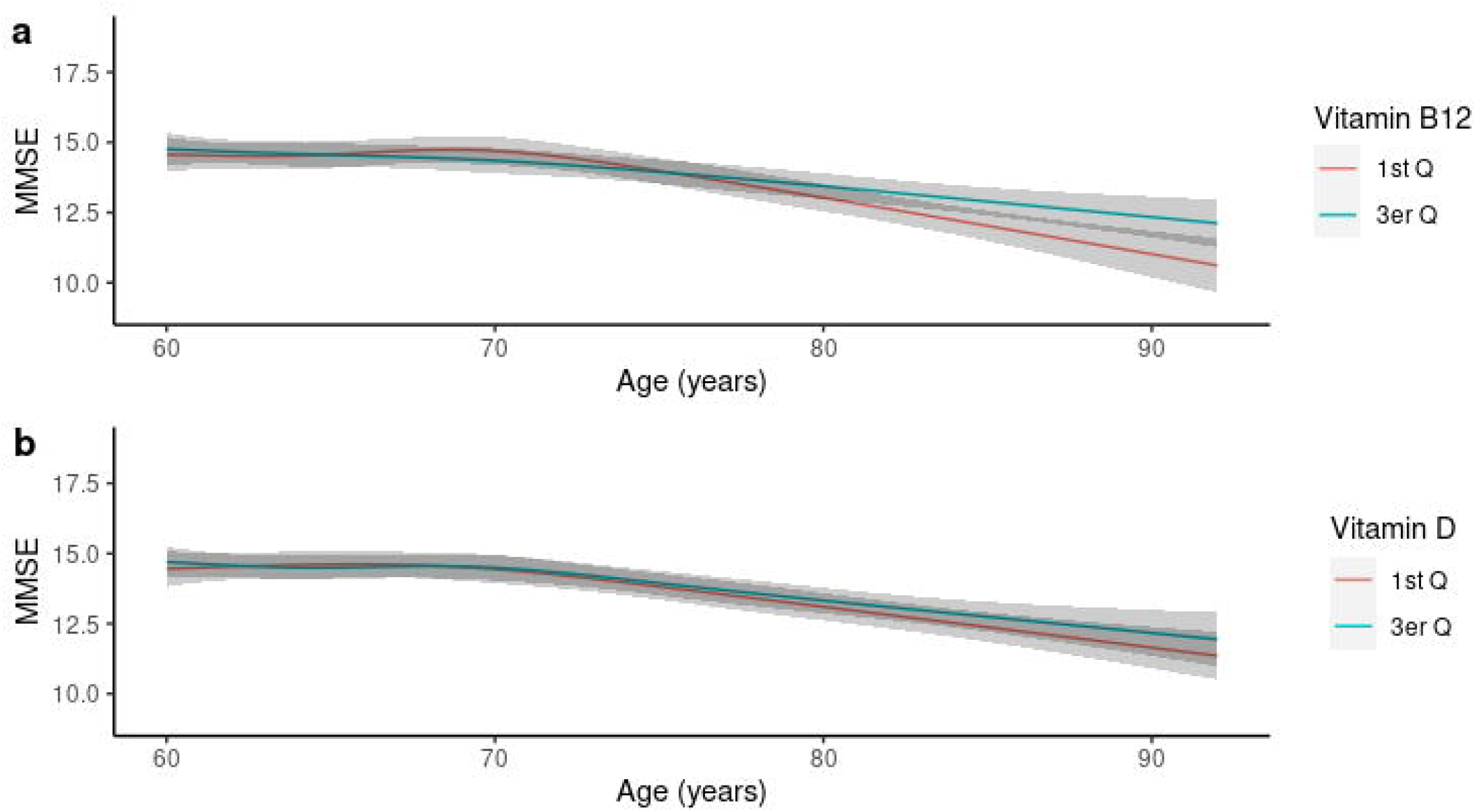

### 3.3 Cognitive function and other covariates

Cognitive function was statically significantly associated with age [F(8) = 17.19, *p* < 0.001], sex [F(1) = 10.68, *p* = 0.001], ethnicity [F(3) = 15.83, *p* < 0.001], years of education [F(3) = 20.09, *p* < 0.001], region [F(3) = 5.94, *p* = 0.001], but not with socioeconomic status [F(4) = 1.38, *p* = 0.240]. In this way, cognitive function decreased with age, was lower in females in comparison to males, was lower in indigenous people in comparison with the other ethnic groups, was higher as years of education increased, was lower for elders residing in urban and rural highland regions in comparison to urban and rural coast regions. Adjusted means of MMSE scores for different covariates by sex are showed in Table 3.

**Table 3.** Adjusted predicted means for MMSE scores. Sample included 1733 individuals. Abbreviations: M, mean; LCI, lower confidence interval; UCI, upper confidence interval; SEE, socioeconomic status

|  |  | Female |  |  | Male |  |  |
| --- | --- | --- | --- | --- | --- | --- | --- |
|  |  | M | LCI | UCI | M | LCI | UCI |
| Age (years) | 60 | 14.64 | 14.11 | 15.17 | 15.06 | 14.53 | 15.60 |
|  | 70 | 14.56 | 14.11 | 15.01 | 14.99 | 14.54 | 15.44 |
|  | 80 | 13.18 | 12.74 | 13.63 | 13.61 | 13.18 | 14.03 |
|  | 90 | 11.52 | 10.81 | 12.23 | 11.95 | 11.23 | 12.66 |
| Education (years) | 0 | 12.86 | 12.41 | 13.31 | 13.29 | 12.80 | 13.77 |
|  | 6 | 15.59 | 15.15 | 16.04 | 16.02 | 15.57 | 16.47 |
|  | 12 | 16.29 | 15.72 | 16.85 | 16.71 | 16.14 | 17.29 |
|  | 18 | 16.50 | 15.61 | 17.40 | 16.93 | 16.03 | 17.83 |
| Ethnic group | Mestizo | 14.61 | 14.17 | 15.05 | 15.03 | 14.59 | 15.48 |
|  | Indigenous | 13.16 | 12.50 | 13.81 | 13.59 | 12.93 | 14.24 |
|  | White | 13.94 | 13.41 | 14.46 | 14.36 | 13.83 | 14.90 |
|  | Other | 13.96 | 13.39 | 14.53 | 14.39 | 13.82 | 14.96 |
| SES | 1 | 14.61 | 14.17 | 15.05 | 15.03 | 14.59 | 15.48 |
|  | 2 | 14.55 | 14.04 | 15.06 | 14.97 | 14.46 | 15.49 |
|  | 3 | 14.83 | 14.33 | 15.33 | 15.26 | 14.76 | 15.75 |
|  | 4 | 14.91 | 14.42 | 15.41 | 15.34 | 14.84 | 15.84 |
|  | 5 | 15.23 | 14.51 | 15.96 | 15.66 | 14.94 | 16.38 |
| Region | Urban highlands | 14.16 | 13.70 | 14.62 | 14.58 | 14.11 | 15.06 |
|  | Urban coast | 14.61 | 14.17 | 15.05 | 15.03 | 14.59 | 15.48 |
|  | Rural highlands | 14.16 | 13.67 | 14.65 | 14.58 | 14.10 | 15.07 |
|  | Rural coast | 14.91 | 14.40 | 15.42 | 15.33 | 14.84 | 15.83 |

## 4. Discussion

The objective of the present work was to study the association of levels of vitamins B12 and D and cognitive function in a population-based context. Independently from sex, age, years of education, ethnicity, socioeconomic status and geographical zone of residence, we found that vitamin B12 but not vitamin D was associated with cognitive function in a representative group of Ecuadorian elders. Elders with lower levels of vitamin B12 showed worse cognitive function that is, lower scores in the MMSE. Interestingly, the drop of cognitive function was particularly steep when elders were 75 years old or older.

### 4.1 Vitamin B12 and D status among Ecuadorian elders

In comparison to international guidelines median levels of B12 among elders of the sample were normal, not so those of vitamin D, which were lower than recommended (see Table 2). Still, 38.49% of elders showed vitamin B12 deficiency. Owing the high prevalence of atrophic gastritis among elders in comparison to the younger population, vitamin B12 deficiency may result from decreased absorption of protein-bound vitamin B12.(22) Atrophic gastritis may also induce reduced release of free vitamin B12 from food proteins and often results in excess of bacteria that consumes nutrients including B12.(22) Although atrophic gastritis is a known risk factor for low vitamin B12 status, it does not fully explain the prevalence of low serum vitamin B12. For instance, it has been described that vitamin B12 deficiency is also common among populations with low prevalence of atrophic gastritis.(23) Other causes of vitamin B12 deficiency among elders may include lack of B12 supplementation or poor consumption of fortified foods, and use of medication such as metformin, omeprazole, etc.(22,24)

Regarding vitamin D, we found that 68.89% of elders showed vitamin D deficiency or insufficiency. Due to methodological discrepancies in relation to the cut-off points for optimal vitamin D values, prevalence of vitamin D deficiency or insufficiency worldwide ranges between extreme values, thus making it difficult to do any comparison.(25) Still, our findings are in agreement with those of Argentine and Brazil, both of which reported a prevalence of vitamin D deficiency or insufficiency of 55% and 87%, respectively.(26,27) Since Ecuador receives sunlight all year around and ultraviolet radiation is necessary for the synthesis of vitamin D, the high prevalence of vitamin D insufficiency and deficiency found in our sample is striking. Vitamin D insufficiency or deficiency in the elderly may arise from the lower amount of ultraviolet radiation received because of the short time elders spent in outdoor activities as well as because the use of sun blockers or other measures to avoid sun exposure.(28,29) Vitamin D insufficiency and deficiency may also arise from a poor daily intake of vitamin D in the diet. In this way, it has been reported that Ecuadorian adults only consume 36% of the recommended daily intake of vitamin D and other nutrients including also vitamin B12.(30)

### 4.2 Vitamin B12 and cognitive function

When evaluating the association between serum levels of vitamin B12 and cognitive function we found that lower levels of vitamin B12 were associated with lower cognitive function. Similar results were reported in a meta-analysis that included data on patients with Parkinson’s disease. (31) This study showed that Parkinson’s disease patients with cognitive dysfunction are more likely to have higher homocysteine levels and lower folate and vitamin B12 levels than patients with normal cognitive function.(31) In contrast, evidence in community-dwelling elders suggests that neither vitamin B12, nor vitamin B6 nor folate show significant benefit on cognition and dementia risk.(32) Furthermore, there is only limited evidence that support the use of vitamin B12 or other vitamin or mineral supplementation to prevent, delay or treat cognitive impairment or dementia. (33–35) Moreover, no evidence has been found suggesting beneficial effects on cognition of supplementation with B vitamins during six to 24 months, neither for lowering homocysteine using B vitamins.(33,35,36)

### 4.3 Vitamin D and cognitive function

We did not found a statistically significantly relationship between vitamin D levels and cognitive function. Our results are in agreement with those of a mendelian randomisation study that found no evidence for serum vitamin D levels as a causal factor for cognitive performance in mid to later life.(13) In contrast, Goodwill and Szoeke found that although evidence based on observational studies demonstrates that low vitamin D is related to poorer cognition, clinical trials are yet to show a clear benefit from vitamin D supplementation.(10) Such conflicting results are probably explained by the quality of available evidence, which in general is not optimal. (37) Vitamin D deficiency or insufficiency are associated with a number of chronic conditions that can precipitate the progression of cognitive decline.(13) Therefore it is plausible that the potential cognitive benefits of vitamin D identified in observational studies may be mediated by improvements in accompanying chronic diseases.(13)

### 4.4 Implications, strengths and limitations

The fact that the use of vitamin B12 supplementation is not effective to prevent, delay or treat cognitive impairment or dementia suggests that foods rich in vitamin B12 cannot be substituted with supplements. Moreover, it seems that once vitamin B12 deficiency is established, damage in cognitive function can no longer be treated or reversed as changes that predispose to cognitive decline probably start early in life.(38,39) Available evidence from randomized controlled trials, which show no obvious cognitive benefit from the use of B12 vitamin supplements, can not be taken as definitive. For instance, most of these studies tend to be too short as to allow to evidence changes in the cognitive function. As existing trials vary greatly in the type of supplementation, the targeted population, the quality of the study and the duration of the treatment, they must necessarily be replicated.

To understand the pathogenic relationship between insufficiency of micronutrients and minerals such as vitamin B12, vitamin D and cognitive function is important to improve quality of life of elders. In Ecuador, as far as we know, a similar study as ours has not been carried out and therefore, our study opens channels of discussion on this regard. According to Guitron, in Ecuador there is a lack of information to estimate the burden of vitamin B12 deficiency specially for vulnerable groups such as the elderly.(41) One of the main limitations of the present study is that with the current design and data is not possible to establish a causal relationship between vitamin B12, vitamin D and cognitive function. Future intervention trials including large, population-diverse populations are needed to assess whether vitamin B12 supplementation can improve cognition in the long-term, even in the absence of clinically manifested vitamin B12 deficiency.(40)

## 5. Conclusions

Low levels of vitamin B12 but not of vitamin D are associated with low cognitive functioning.

## Data Availability

Data is publicly available and can be accessed via https://anda.inec.gob.ec/anda/index.php/catalog/292

## Acknowledgments

We would like to thank to Arq. Carlos Martín Román, Prof.dr. Chris de Zeeuw, Dr. María Paulina Robalino Valdivieso for their academic support.

## Author contribution

The main author of this work declares that all authors have contributed and work on the development of it as follows. AFVV: Conception and design, analysis and interpretation of data, drafting of the article, final approval of the version to be published. TVCA: Conception and design, analysis and interpretation of data, drafting of the article, final approval of the version to be published. JSVM Conception and design, analysis and interpretation of data, drafting of the article, final approval of the version to be published. ECTV: Critical revision for important intellectual content, writing and final approval of the version to be published. JSPA: Critical revision for important intellectual content, writing and final approval of the version to be published. TMNC: Critical revision for important intellectual content, writing and final approval of the version to be published. SIHA: Critical revision for important intellectual content, writing and final approval of the version to be published. MFVV: Conception and design, analysis and interpretation of data, drafting of the article, critical revision for important intellectual content, final approval of the version to be published.

