## Supplementary figures and images for "Cognitive function and vitamin B12 and D in elders from Ecuador"

### Supplementary figure 1

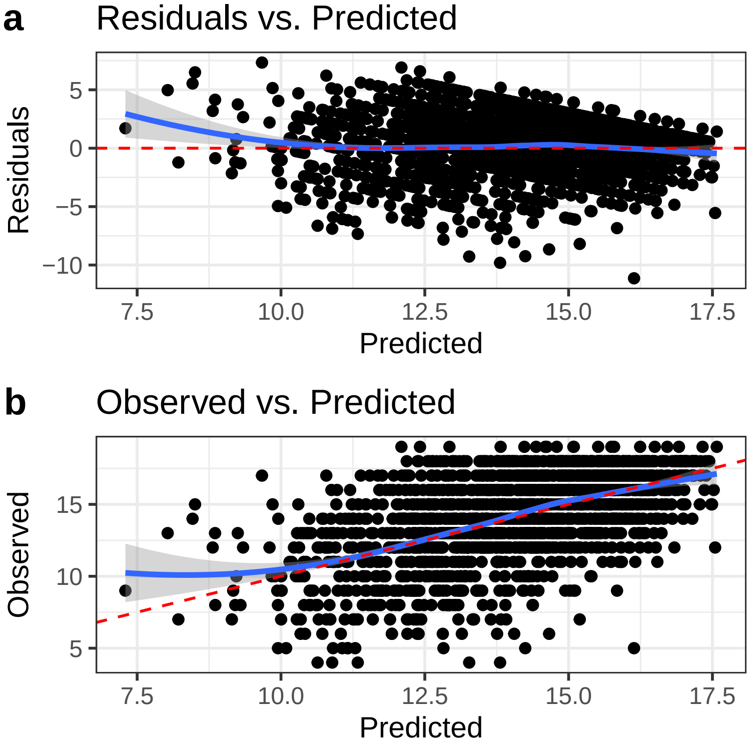
